# Head-to-head comparison of the RMI and ADNEX models to estimate the risk of ovarian malignancy: systematic review and meta-analysis of external validation studies

**DOI:** 10.1101/2024.11.29.24318146

**Authors:** Lasai Barreñada, Ashleigh Ledger, Agnieszka Kotlarz, Paula Dhiman, Gary S. Collins, Laure Wynants, Jan Y. Verbakel, Lil Valentin, Dirk Timmerman, Ben Van Calster

**Affiliations:** Department of Development and Regeneration, KU Leuven, Belgium; Department of Gynecology and Oncology, Faculty of Medicine, Jagiellonian University Medical College, Krakow, Poland; Centre for Statistics in Medicine, Nuffield Department of Orthopaedics, Rheumatology and Musculoskeletal Sciences, University of Oxford, Oxford OX3 7LD, UK; Department of Epidemiology, CAPHRI Care and Public Health Research Institute, Maastricht University, Maastricht, Netherlands.; Department of Public Health and Primary Care, KU Leuven, Belgium; Leuven Unit for Health Technology Assessment Research (LUHTAR), KU Leuven, Belgium; Department of Obstetrics and Gynaecology, University Hospitals Leuven, Leuven, Belgium; Department of Obstetrics and Gynaecology, Skåne University Hospital, Malmö, Sweden; Department of Clinical Sciences Malmö, Lund University, Lund, Sweden

## Abstract

**Background:** ADNEX and RMI are models to estimate the risk of malignancy of ovarian masses based on clinical and ultrasound information. The aim of this systematic review and meta-analysis is to synthesise head to-head comparisons of these models.

**Methods:** We performed a systematic literature search up to 31/07/2024. We included all external validation studies of the performance of ADNEX and RMI on the same data. We did a random effects meta-analysis of the area under the receiver operating characteristic curve (AUC), sensitivity, specificity, net benefit and relative utility at 10% malignancy risk threshold for ADNEX and 200 cutoff for RMI.

**Results:** We included 11 studies comprising 8271 tumours. Most studies were at high risk of bias (incomplete reporting, poor methodology). For ADNEX with CA125 vs RMI, the summary AUC to distinguish benign from malignant tumours in operated patients was 0.92 (CI 0.90-0.94) for ADNEX and 0.85 (CI 0.80-0.89) for RMI. Sensitivity and specificity for ADNEX were 0.93 (0.90-0.96) and 0.77 (0.71-0.81). For RMI they were 0.61 (0.56-0.67) and 0.93 (0.90-0.95). The probability of ADNEX being clinically useful in operated patients was 96% vs 15% for RMI at the selected cutoffs (10%, 200).

**Conclusion:** ADNEX is clinically more useful than RMI.

**Systematic review registration:** CRD42023449454

## BACKGROUND

To choose the optimal management of an ovarian mass, the characterisation of the mass as benign or malignant is essential (1,2). If the mass is likely to be malignant, treatment in a referral centre for gynaecological oncology improves outcome (3,4). Advanced imaging and predictive models have greatly improved our ability to distinguish between benign and malignant ovarian masses before surgery. This should result in better treatment outcomes. The Assessment of Different NEoplasias in the adneXa (ADNEX) model (5) and the Risk of Malignancy Index (RMI) (6) have become key tools for estimating the risk of an ovarian mass being malignant.

RMI is an index developed in 1990 that provides a value between 0 and infinity, where higher values are associated with malignancy (6). A commonly used cutoff to classify a tumour as high risk for malignancy is 200 (7–11), but other cutoffs have also been suggested (7,12,13). RMI is calculated as the product of three variables: the CA125 serum level (U/ml), ultrasound score (0,1 or 3), and menopausal status (1 premenopausal; 3 postmenopausal). When calculating the ultrasound score, 1 point is given for each of the following ultrasound features: multilocular cyst, solid areas, evidence of metastases, ascites and bilaterality. If none of the ultrasound features is present the score is 0, and therefore RMI is 0. If one ultrasound feature is present the score is 1, and if more than one ultrasound feature is present the score is 3. The model was developed using data from 143 patients of whom 42 had a malignant tumour (malignancy proportion 29%). On internal validation, the sensitivity and specificity were 85% and 97%, respectively, at the 200 RMI score cutoff (6). On external validation, the sensitivity and specificity at the 200 cutoff were 60.4% and 95.3%, respectively (14). Modifications of the RMI have been published (RMI 2, RMI 3 and, RMI 4 (15–17)), but in several countries the use of the original RMI is recommended (7–9). ADNEX is a risk prediction model published in 2014. It estimates the risk that an ovarian tumour is benign, borderline, stage I primary invasive, stage II-IV primary invasive or a metastasis in the ovary from another primary tumour (5). The estimated risk of malignancy is calculated as the sum of the risks of the four malignant subtypes. ADNEX is a multinomial logistic regression model that uses nine predictors: patient’s age (years), maximum diameter of lesion (mm), proportion of solid tissue (calculated as the maximum diameter of the largest solid component divided by the maximum diameter of the lesion), number of papillary projections (ordinal), presence of acoustic shadows, presence of ascites, presence of more than 10 cyst locules, and type of centre (oncology centre vs other), and serum CA125 (U/ml)). There is also an ADNEX version without serum CA125. ADNEX was developed on data from 3506 patients and temporally validated on data from 2403 patients. The final published formula was based on retraining the model on the combined data from all 5909 patients. On internal validation using the combined data the area under the receiver operating characteristic curve (AUC) of ADNEX with CA125 was 0.95, and using a risk of malignancy cutoff of 10%, the cutoff recommended in an international consensus statement (1), the sensitivity was 96.4% and the specificity 73.2% (1). To the best of our knowledge there are only two published meta-analyses presenting results of a head-to head comparison of ADNEX with RMI. One compared sensitivity and specificity at the 10% risk of malignancy cutoff for ADNEX and for the 200 cutoff for RMI and found the sensitivity to be higher and the specificity lower for ADNEX (18). Another more comprehensive meta-analysis presents results for classification, discrimination and clinical utility. It includes the results from 17 centres participating in the same international multicentre study and showed ADNEX to be superior to RMI (14). In some national guidelines, RMI is still recommended for triaging patients with ovarian tumours for referral to an oncology centre (7–11,13). Therefore, an extended head-to-head comparison of the performance of RMI and ADNEX is needed.

The aim of this work is to present a systematic review and meta-analysis of studies that externally validated ADNEX and the original RMI (RMI 1) on the same data (head-to head comparison).

## METHODS

### Protocol registration

We report this study according to the PRISMA (Preferred Reporting Items for Systematic Reviews and Meta-Analysis) and TRIPOD-SRMA (Transparent Reporting of multivariable prediction models for Individual Prognosis Or Diagnosis: checklist for Systematic Reviews and Meta-Analysis) checklists (19,20). The study protocol was prospectively registered in the international prospective register of systematic reviews (PROSPERO; ID CRD42023449454).

### Eligibility criteria

This is a follow-up study of a systematic review of ADNEX (21). Inclusion and exclusion criteria are the same as in the systematic review of ADNEX, with the restriction that eligible studies also had to present metrics for RMI 1 at the 200 cutoff for the same study population. The 200 cutoff was selected as it is a cutoff often recommended for referring a patient to an oncological centre (7–11).

The inclusion criteria were: any publication that externally validates the performance of the ADNEX model and RMI simultaneously on the same study population using any study design and any study population consisting of patients with an adnexal mass.

The exclusion criteria were: (1) studies that do not evaluate the predictive performance (in any way) of ADNEX and RMI simultaneously in the same population, (2) studies that only evaluate the predictive performance of updated versions of ADNEX or RMI. Updating can refer to recalibration, refitting, extension with additional predictors, or any type of modification to the original formula. Thus, RMI 2, 3 or 4 or any updated ADNEX model are not included in this review. (3) Studies for which only an abstract is available, either because we could not get access to the full text, or because no full text exists (e.g. conference abstracts) and (4) case studies presenting the performance in individual patients. When the full text was not accessible, we tried to obtain it by contacting the corresponding authors via email.

### Information sources and search strategy

The search was conducted on the 31^st^ of July 2024. The search string and overall search strategy were created with the help of biomedical librarians at the KU Leuven Libraries. Embase, Web of Science, Scopus, and Medline (via PubMed) were searched for published studies, and EuropePMC for preprints. The databases were searched from the publication of the first ADNEX article (15/10/2014) until 31/07/2024. To retrieve any additional publications, we performed forward and backward snowballing of the included articles (looking through citations and references of the original article), checked the reference lists of relevant reviews and opinion articles, and screened all articles citing the original ADNEX publication. No restriction was placed on language, but for papers written in languages other than English, Spanish, Dutch, French, or Swedish, we used an automated translation tool (deepl.com) to decide on paper inclusion or exclusion and to extract information. The full search strategy is provided in **Supplementary Material S1**.

### Study selection

The studies identified in our search were imported into Zotero reference manager, where they were automatically deduplicated. The deduplicated records were then imported into the Rayyan web application for further manual deduplication (by LB) and subsequent screening of the title and abstract by two independent authors (LB and AL). Disagreements in eligibility were resolved by discussion between LB and AL.

Three of the authors (BVC, LV, and DT) were members of the IOTA group that developed ADNEX. Therefore, we divided the studies into those that were linked or not linked to IOTA. A study was linked to IOTA if it was coauthored by a member of the IOTA steering committee. IOTA linked papers, as well as a few others with a potential conflict of interest (i.e., including authors that are or were IOTA collaborators), were assessed by two authors that are independent from IOTA (PD and GSC, medical statisticians with expertise in prediction modelling). All other studies were independently assessed by LB and AL. Disagreements were resolved by discussion between reviewers (pair of LB-AL or GSC-PD), and for the non-IOTA papers, by discussion with authors BVC, LV, and JYV.

### Data extraction and data items

Information was collected and entered into a standardised data extraction form in Microsoft Excel by LB, AL, PD, and GSC. The extraction process focused on study design, target population, reference standards, sample size, performance results, completeness of reporting, quality of methodology, and risk of bias (**Supplementary Table S1**). The extraction form was structured based on CHARMS (Checklist for critical Appraisal and data extraction for systematic reviews of prediction Modelling Studies), TRIPOD (Transparent Reporting of a multivariable prediction model for Individual Prognosis Or Diagnosis), and PROBAST (Prediction model Risk of Bias Assessment Tool) tools (22–24).

To describe the performance of ADNEX, we collected information regarding discrimination (AUC), calibration (calibration slope, calibration intercept, calibration curve), clinical utility (net benefit) and classification performance (sensitivity, specificity) at the 10% risk of malignancy threshold. We also collected information on the AUC, sensitivity, and specificity at the 200 cutoff for RMI. When a performance measure was missing or the cutoff was not clear, we contacted the authors for clarification.

For each study, we evaluated the reporting of all relevant TRIPOD items for external validation studies. We also applied the PROBAST signalling questions and assessed the risk of bias within each domain (participants, predictors, outcome, and analysis), as well as the overall risk of bias. The overall risk of bias is determined by the worst risk of bias rating across the domains. We included an explanation for our classification of the risk of bias. We critically appraised ADNEX and RMI independently because their predictors differ, the development information differs, and the relevant performance measures differ. However, most signalling questions of PROBAST and most TRIPOD items are assessed at study level and are therefore equal for the two models.

### Statistical analysis and quantitative data synthesis

We performed a meta-analysis of centre specific results (or study specific when results for individual centres were not available) using random effects meta-analysis methods. The AUC, sensitivity and specificity were meta-analysed on the logit scale. We calculated 95% confidence intervals (CI) for the summary estimates using Hartung-Knapp-Sidik-Jonkman (HKSJ) method, and estimated the between study heterogeneity using tau squared and 95% prediction intervals (PI)(25,26).Unreported confidence intervals and prediction intervals were calculated based on Debray et al (2019) for AUC and with Wilson’s method for specificity and sensitivity (27,28).

We express clinical utility to decide which patients to refer to an oncology centre as net benefit (NB), relative utility (RU) and P-useful (29–33). The higher NB, RU and P-useful, the better. NB combines the benefits of true positives and the harms of false positives on a single scale by using a weighting factor for false positives. This weighting factor corresponds to the odds of the chosen risk threshold T [i.e., T/(1-T)] to select patients for treatment. We used a risk threshold of 10%. This means that we considered the benefit of referring a patient with an adnexal malignancy to an oncology centre to be 9 times as large as the harm of referring one patient with a benign tumor to an oncology centre. To evaluate clinical utility we performed a trivariate meta-analysis of sensitivity, specificity, and prevalence of malignancy for both ADNEX and RMI. Because RMI does not provide risk estimates, we used a cutoff of 200 and calculated NB at the 10% threshold. P-useful measures the probability that a model is useful in a new centre (NB difference with best default > 0). For NB and RU, we report 95% credible intervals (CrI) instead of CI. See **Supplementary material S2 and S3** for more information on the methodology.

Four meta-analyses were performed. We compared ADNEX with CA125 with RMI and ADNEX without CA125 with RMI in two populations: 1) only operated patients, and 2) operated and conservatively managed patients combined.

The analyses were performed in R version 4.4.0 using package “metamisc” for AUC, “meta” and “mada” for sensitivity and specificity, and rjags for NB and RU. Bayesian methods for meta-analysis of clinical utility were computed using JAGS version 4.3.0.

## RESULTS

Thirteen studies presented results for both ADNEX and RMI (**Fig. S1, Table S3**)(14,34–45). Two studies were excluded from the systematic review and the meta-analysis because they validated only RMI 2(42,45).

The characteristics of the included studies are summarised in **Table 1**. The total number of patients included in the systematic review is 8271 with a median sample size of 326 patients per study (range 100-4905). The studies were conducted in 14 countries. The studies conducted 11 validations of RMI and 15 validations of ADNEX, 11 for the version with CA125 and 4 for the version without CA125. The complete extraction data and code to reproduce the results and figures is available in the OSF repository (https://osf.io/nt89z/)

**Table 1.** Characteristics of the eleven included studies.

| Study characteristics | N (%) | Comments |
| --- | --- | --- |
| Unit of analysis |  |  |
| Patient | 10 (91) |  |
| Tumour | 1 (9) |  |
| Region |  |  |
| Asia | 6 (55) | Includes two studies from Turkey |
| Europe | 4 (36) |  |
| South America | 1 (9) |  |
| Number of centres |  |  |
| 1 | 7 (64) |  |
| 2-5 | 3 (27) |  |
| >5 | 1 (9) |  |
| Type of centre |  |  |
| Oncology centre(s) | 8 (73) |  |
| Non-oncology centre(s) | 0 |  |
| Both types of centres | 1 (9) |  |
| Unclear | 2 (18) |  |
| Type of study |  |  |
| Prospective | 4 (36) |  |
| Retrospective | 5 (45) |  |
| Unclear | 2 (18) |  |
| ADNEX version |  |  |
| With CA125 | 11 (100) | 7 only used ADNEX with CA125, 4 used both |
| Without CA125 | 4 (36) |  |
| Target population |  |  |
| Surgically managed patients | 10 (91) |  |
| Surgically and non-surgically managed patients | 1 (9) |  |

### Critical appraisal: reporting and risk of bias

The eleven studies reported on average 19 out of 28 (68%) TRIPOD items for the validation of ADNEX, and 17.18 (61%) TRIPOD items for the validation of RMI **(Fig. S2, Table S4)**. The items that were reported in less than 35% of the validations both for ADNEX and RMI were: Justification of sample size (item 8) and justification of missing data management (item 9), specifying relevant measures of performance (item 10d) with confidence intervals (item 16) and comparing the participantś characteristics in the original data with those in the validation data and discussing any differences (item 13c).

Based on PROBAST, 12 of 15 validations (80%) of ADNEX were rated as overall high risk of bias, while nine out of 11 validations (82%) of RMI were rated as overall high risk of bias mainly due to the small sample size and the exclusion of participants with missing data (**Fig. 1 and Figs. S3-4**). Most validations were unclear in the predictors domain due to not specifying when in relation to the inclusion scan serum levels of CA125 were measured, or for not reporting if the assessment of predictors was blinded to the outcome.

**Fig. 1:**
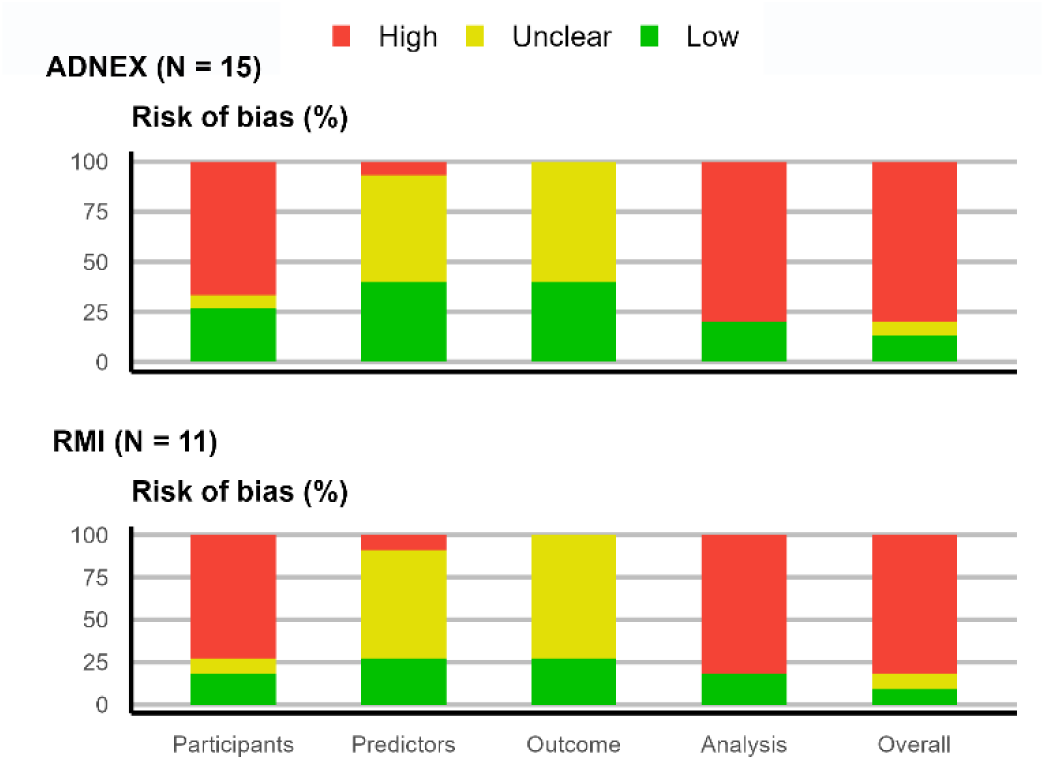
Risk of bias by subdomain and overall for Assessment of Different NEoplasias in the adneXa (ADNEX) and Risk of malignancy index (RMI) validations using the Prediction model Risk Of Bias ASsessment Tool (PROBAST). There are 15 validations of ADNEX and 11 validations of RMI.

### Meta-analysis inclusions

One study was excluded from the meta-analysis because it only presented results stratified by the certainty of the ultrasound examiner when assessing the outcome (40) and another study was excluded for not reporting the cutoff used to calculate the sensitivity and specificity of ADNEX (44). Therefore, 9 studies where eligible for meta-analysis (14,34–39,41,43). Four studies presented results for both ADNEX versions (with and without CA125) and five presented results only for ADNEX with CA125. AUC results were reported in all studies except one (39). However, one study presented AUC calculated using predicted outcome instead of predicted probability (ROC curve based on one single point) and thus was excluded from meta-analysis of AUC (41). Therefore, the meta-analysis of AUC was based on seven studies (14,34–38,43). Sensitivity and Specificity at the 10% risk of malignancy threshold for ADNEX was available (either in the publication or after contacting the authors) for all studies except one (36) (**Table S4**).

### Meta-analyses of AUC

Results of the meta-analysis of AUC are shown in **Table 2** and **Figures S5-S6**. Seven studies included in the meta-analysis compared the AUC of ADNEX with CA125 with that of RMI in operated patients (14,34–38,43). The summary estimate for ADNEX was 0.92 and that for RMI was 0.85 (**Table 2, Fig. S5**). Four studies included in the meta-analysis compared the AUC of ADNEX without CA125 with that of RMI (14,34,38,43). The summary estimate for ADNEX was 0.92, that for RMI was 0.87 (**Table 2, Fig. S6**). Only one multicentre study (14) reported results for both operated and conservatively managed patients with a summary estimate of 0.93 for ADNEX with CA125 and 0.89 for RMI. The corresponding summary estimate for ADNEX without CA125 was 0.94 versus 0.89 for RMI (**Table 2, Table S4**). **Figure 2** shows the centre specific or study specific comparison of ADNEX with or without CA125 vs RMI in only operated patients.

**Fig. 2:**
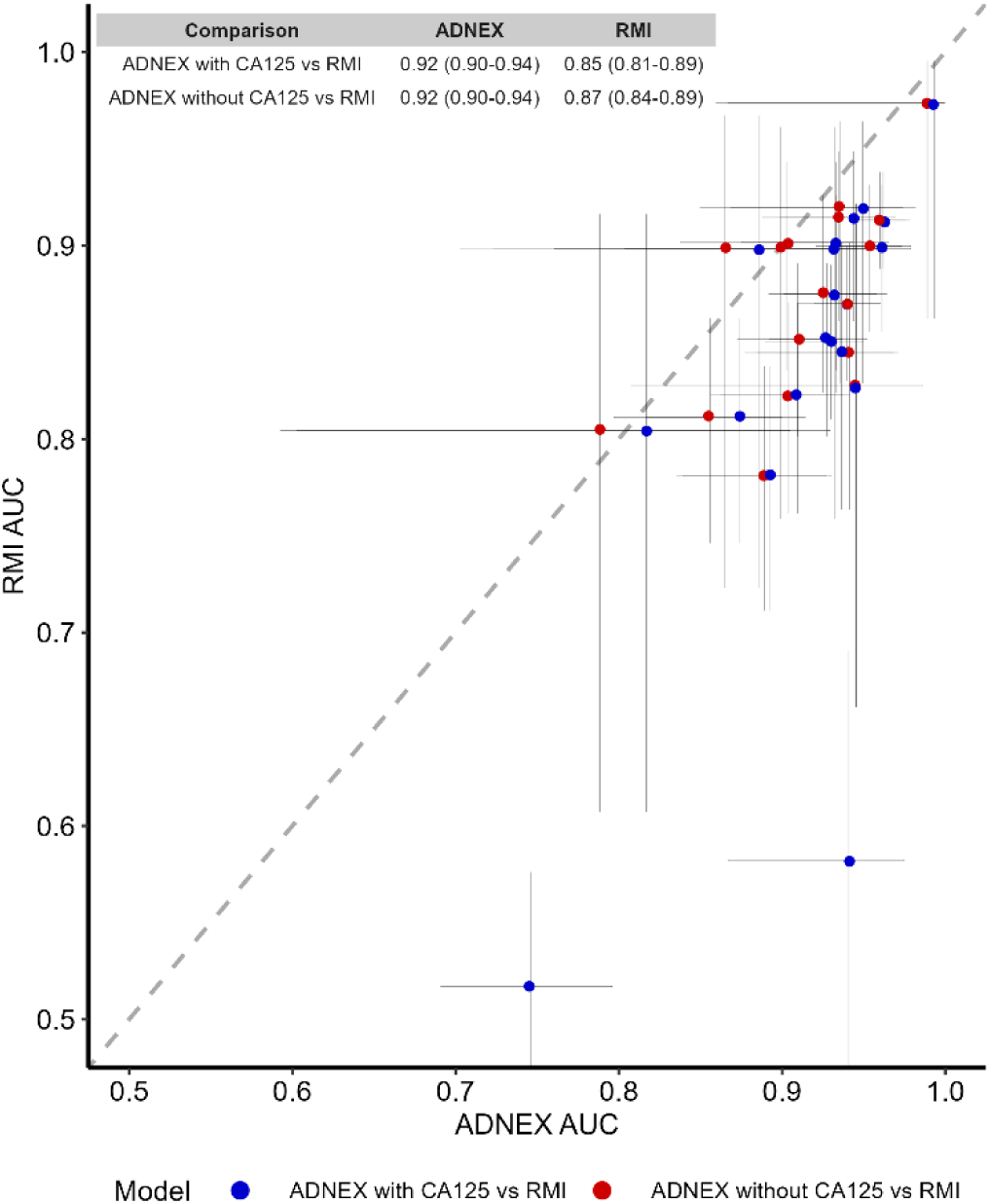
Comparison of the area under the receiver operating characteristic curve (AUC) of ADNEX (with and without CA125) and RMI per centre in the included studies for only operated patients. Vertical and horizontal lines present the confidence interval of each centre’s AUC. Dots below the diagonal line indicate higher AUC for ADNEX than for RMI. Dots above the diagonal line indicate higher AUC for RMI.

**Table 2.** Meta analysis results for the area under the receiver operating characteristic curve (AUC).

| Meta-analysis | Patients | Studies | Centres | Summary estimate | 95% CI | 95% PI | $\tau^2$ |
| --- | --- | --- | --- | --- | --- | --- | --- |
| *Target population: operated patients* |  |  |  |  |  |  |  |
| *ADNEX with CA125 vs RMI* |  |  |  |  |  |  |  |
| ADNEX | 4668 | 7 | 26 | 0.92 | 0.90 to 0.94 | 0.82 to 0.99 | 0.30 |
| RMI | 4668 | 7 | 26 | 0.85 | 0.81 to 0.89 | 0.64 to 0.98 | 0.44 |
| *ADNEX without CA125 vs RMI* |  |  |  |  |  |  |  |
| ADNEX | 3773 | 4 | 22 | 0.92 | 0.90 to 0.94 | 0.85 to 0.97 | 0.16 |
| RMI | 3773 | 4 | 22 | 0.87 | 0.84 to 0.89 | 0.78 to 0.94 | 0.12 |
| *Target population: Operated and conservatively managed patients* |  |  |  |  |  |  |  |
| *ADNEX with CA125 vs RMI* |  |  |  |  |  |  |  |
| ADNEX | 4905 | 1 | 17 | 0.93 | 0.91 to 0.96 | 0.85 to 0.99 | 0.27 |
| RMI | 4905 | 1 | 17 | 0.89 | 0.86 to 0.92 | 0.77 to 0.97 | 0.22 |
| *ADNEX without CA125 vs RMI* |  |  |  |  |  |  |  |
| ADNEX | 4905 | 1 | 17 | 0.94 | 0.93 to 0.96 | 0.88 to 0.99 | 0.26 |
| RMI | 4905 | 1 | 17 | 0.89 | 0.86 to 0.92 | 0.77 to 0.97 | 0.22 |

CI, confidence interval; PI, prediction interval; AUC, Area under the receiving operating characteristic curve; $\tau^2$ , tau squared, RMI, Risk of malignancy index; ADNEX, Assessment of Different NEoplasias in the adneXa

### Meta-analyses of sensitivity and specificity

The results of the meta-analysis of sensitivity and specificity are presented in **Table 3** and **Figures S7-S8**. Eight studies included in the meta-analysis compared the sensitivity and specificity of ADNEX with CA125 with those of RMI in surgically managed patients (14,34,35,37–39,41,43). ADNEX had a sensitivity (summary estimate) of 0.93 and a specificity (summary estimate) of 0.77, while RMI had a sensitivity of 0.61 and a specificity of 0.92 (**Table 3, Fig. S7**). Four studies included in the meta-analysis compared the sensitivity and specificity of ADNEX without CA125 with that of RMI in surgically managed patients (14,34,38,43). In these studies, ADNEX had sensitivity 0.94 and specificity 0.76, while RMI had sensitivity 0.61 and specificity 0.93 (**Table 3, Fig. S8**). **Figure 3** illustrates the centre-specific sensitivities and specificities of ADNEX and RMI in patients managed with surgery in the space of a receiver operating characteristic curve. Only one multicentre study (14) reported results for both operated and conservatively managed patients. In that study, the summary estimate of sensitivity for ADNEX with or without CA125 was 0.92 and that of specificity was 0.85. For RMI the corresponding summary estimates were 0.61 and 0.95 (**Table 3**).

**Fig. 3:**
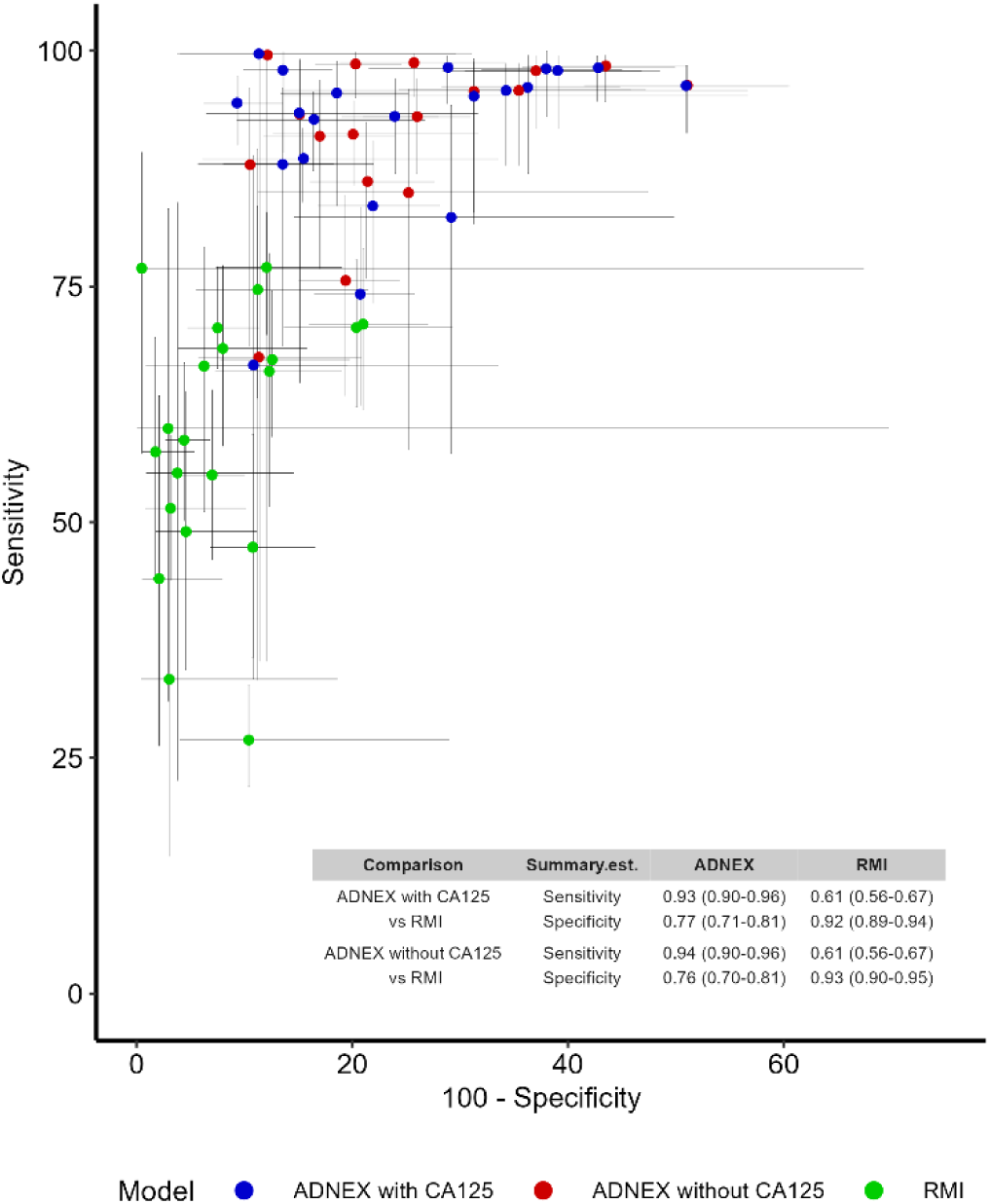
Sensitivity and specificity of ADNEX (10% cutoff) and RMI (cutoff 200) in surgically managed patients illustrated in the space of a receiver operating characteristic curve. Each point represents the result of one study or centre and the lines represent the 95% confidence interval.

**Table 3.**
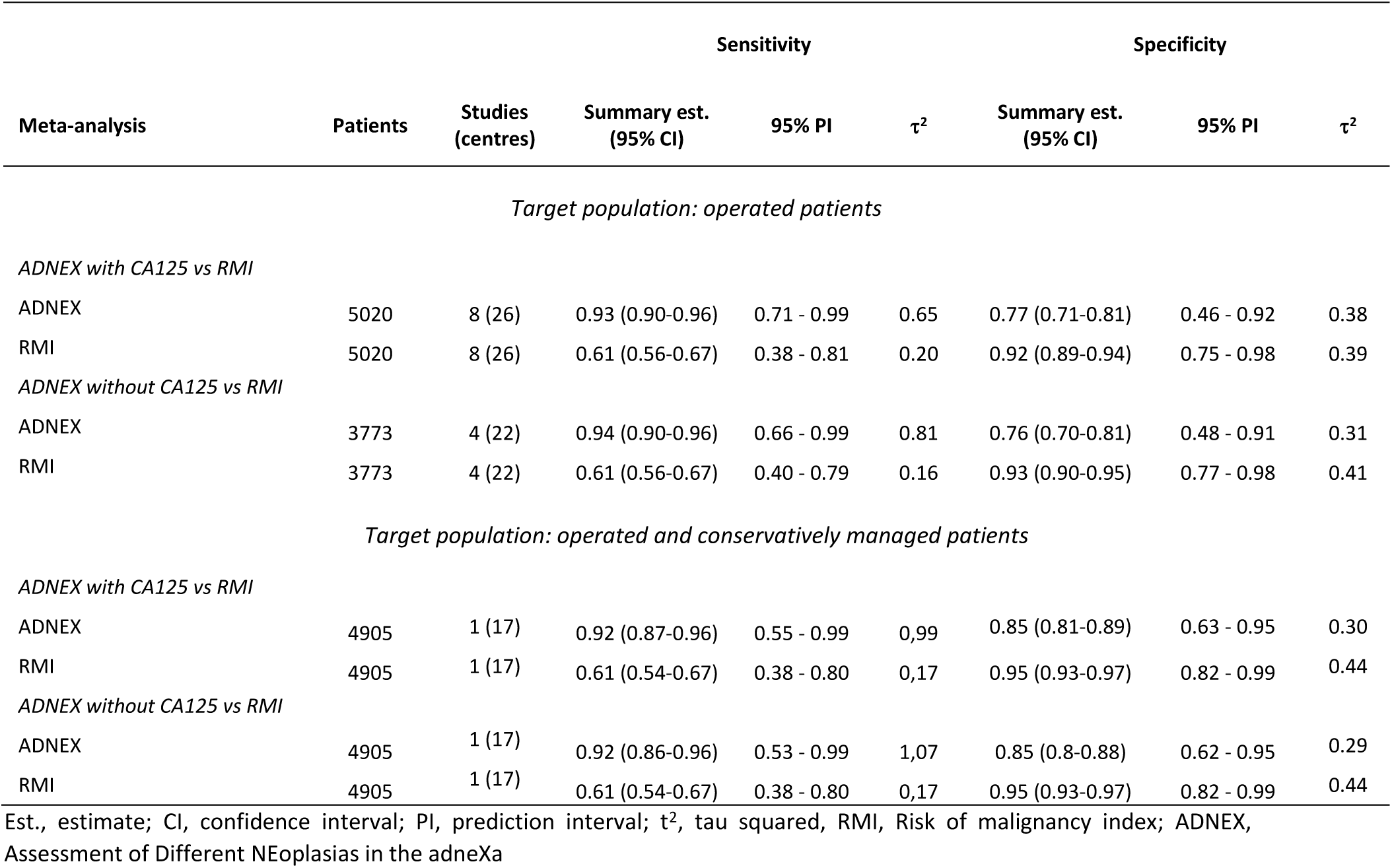
Meta analysis results for sensitivity and specificity at the 10% risk of malignancy cutoff for ADNEX and at the 200 cutoff for RMI.

| Meta-analysis | Patients | Studies<br>(centres) | Sensitivity |  |  | Specificity |  |  |
| --- | --- | --- | --- | --- | --- | --- | --- | --- |
| | | | Summary est.<br>(95% CI) | 95% PI | $\tau^2$ | Summary est.<br>(95% CI) | 95% PI | $\tau^2$ |
| Target population: operated patients |  |  |  |  |  |  |  |  |
| ADNEX with CA125 vs RMI |  |  |  |  |  |  |  |  |
| ADNEX | 5020 | 8 (26) | 0.93 (0.90-0.96) | 0.71 - 0.99 | 0.65 | 0.77 (0.71-0.81) | 0.46 - 0.92 | 0.38 |
| RMI | 5020 | 8 (26) | 0.61 (0.56-0.67) | 0.38 - 0.81 | 0.20 | 0.92 (0.89-0.94) | 0.75 - 0.98 | 0.39 |
| ADNEX without CA125 vs RMI |  |  |  |  |  |  |  |  |
| ADNEX | 3773 | 4 (22) | 0.94 (0.90-0.96) | 0.66 - 0.99 | 0.81 | 0.76 (0.70-0.81) | 0.48 - 0.91 | 0.31 |
| RMI | 3773 | 4 (22) | 0.61 (0.56-0.67) | 0.40 - 0.79 | 0.16 | 0.93 (0.90-0.95) | 0.77 - 0.98 | 0.41 |
| Target population: operated and conservatively managed patients |  |  |  |  |  |  |  |  |
| ADNEX with CA125 vs RMI |  |  |  |  |  |  |  |  |
| ADNEX | 4905 | 1 (17) | 0.92 (0.87-0.96) | 0.55 - 0.99 | 0,99 | 0.85 (0.81-0.89) | 0.63 - 0.95 | 0.30 |
| RMI | 4905 | 1 (17) | 0.61 (0.54-0.67) | 0.38 - 0.80 | 0,17 | 0.95 (0.93-0.97) | 0.82 - 0.99 | 0.44 |
| ADNEX without CA125 vs RMI |  |  |  |  |  |  |  |  |
| ADNEX | 4905 | 1 (17) | 0.92 (0.86-0.96) | 0.53 - 0.99 | 1,07 | 0.85 (0.8-0.88) | 0.62 - 0.95 | 0.29 |
| RMI | 4905 | 1 (17) | 0.61 (0.54-0.67) | 0.38 - 0.80 | 0,17 | 0.95 (0.93-0.97) | 0.82 - 0.99 | 0.44 |

Est., estimate; CI, confidence interval; PI, prediction interval; $\tau^2$ , tau squared, RMI, Risk of malignancy index; ADNEX, Assessment of Different NEoplasias in the adneXa

### Meta-analyses of clinical utility

The results for the meta-analysis of clinical utility are shown in **Table 4**. The summary NB for ADNEX with CA125 was 0.29 and that of RMI was 0.19. The RU was 0.51 for ADNEX with CA125 vs -0.80 for RMI. The probability that the model would be clinically useful in a new centre was 96% for ADNEX with CA125 and 15% for RMI. The summary NB for ADNEX without CA125 was 0.30 and that of RMI was 0.20, the corresponding RUs were 0.48 vs -0.90. The probability that the model would be clinically useful in a new centre was 95% for ADNEX without CA 125 and 13% for RMI. In patients either surgically or conservatively managed, ADNEX with CA125 had summary NB 0.17 and RU 0.69, ADNEX without CA125 had summary NB 0.17 and RU 0.68, and RMI had summary NB 0.12 and RU 0.05.

**Table 4.** Meta-analysis for clinical utility of ADNEX and RMI.

| Meta-analysis | Patients | Studies (centres) | Net benefit<br>Summary est.<br>(95% CrI) | 95% PI | Relative utility<br>Summary est.<br>(95% CrI) | 95% PI | P(useful) |
| --- | --- | --- | --- | --- | --- | --- | --- |
| *Target population: operated patients* |  |  |  |  |  |  |  |
| ADNEX with CA125 vs RMI |  |  |  |  |  |  |  |
| ADNEX | 5020 | 8 (26) | 0.29 (0.21-0.37) | 0.05 - 0.70 | 0.51 (0.4-0.61) | -0.11 - 0.77 | 96% |
| RMI | 5020 | 8 (26) | 0.19 (0.13-0.27) | 0.02 - 0.60 | -0.8 (-1.30--0.37) | -0.80 (-1.30 - -0.37) | 15% |
| ADNEX without CA125 vs RMI |  |  |  |  |  |  |  |
| ADNEX | 3773 | 4 (22) | 0.30 (0.22-0.39) | 0.07 - 0.67 | 0.48 (0.34-0.61) | -0.14 - 0.78 | 95% |
| RMI | 3773 | 4 (22) | 0.20 (0.13-0.28) | 0.02 - 0.58 | -0.90 (-1.46--0.45) | -3.97 - 0.31 | 13% |
| *Target population: operated and conservatively managed patients* |  |  |  |  |  |  |  |
| ADNEX with CA125 vs RMI |  |  |  |  |  |  |  |
| ADNEX | 4905 | 1 (17) | 0.17 (0.10-0.26) | 0.02 - 0.61 | 0.69 (0.60-0.77) | 0.20 - 0.85 | 99% |
| RMI | 4905 | 1 (17) | 0.12 (0.06-0.19) | 0.01 - 0.50 | 0.05 (-0.27-0.32) | -1.58 - 0.53 | 58% |
| ADNEX without CA125 vs RMI |  |  |  |  |  |  |  |
| ADNEX | 4905 | 1 (17) | 0.17 (0.11-0.25) | 0.02 - 0.59 | 0.68 (0.58-0.77) | 0.18 - 0.85 | 99% |
| RMI | 4905 | 1 (17) | 0.12 (0.06-0.19) | 0.01 - 0.50 | 0.05 (-0.27-0.32) | -1.58 - 0.53 | 58% |

Est., estimate; CrI; credible interval, PI; prediction interval, RMI; Risk of malignancy index, ADNEX; Assessment of Different NEoplasias in the adneXa

## DISCUSSION

This meta-analysis shows that the ability of ADNEX with or without CA125 to discriminate between benign and malignant adnexal masses is superior to that of RMI. It also shows that that the clinical utility of ADNEX is superior to that of RMI when using the commonly recommended cutoffs (10% malignancy risk and RMI score 200) to decide which patients with an adnexal mass to refer to an oncology centre.

The strength of our meta-analysis is that it is a comprehensive head-to-head to comparison of ADNEX with RMI. Only two published meta-analyses compared the performance of ADNEX and RMI head-to-head (14,18). One included two studies and compared only sensitivity and specificity. The other one is included in the current systematic review (14). A limitation is the high risk of bias in all but one of the included studies, and the poor reporting of TRIPOD items. Even though this is not a limitation of our meta-analysis, it affects our results. Some of the reasons for high risk of bias, e.g. unknown or inappropriate handling of missing data (inappropriate exclusions), unclear information on blinding, and small sample size may have biased the results of the included studies, and therefore also the results of our meta-analysis. On the other hand, the summary estimates for AUC, sensitivity, and specificity in the only study with low risk of bias (14) (a meta-analysis of results from 17 centres) are very similar to those of the studies with high risk of bias. The lower summary net benefit in the study with low risk of bias is explained by the lower prevalence of malignancy in that study, which in turn is explained by it including both surgically and conservatively managed patients. All studies with high risk of bias included only surgically managed patients. Because of absent reporting of calibration performance in all but one of the included studies we could not perform meta-analysis of calibration performance, which is another limitation of our meta-analysis. It was also not possible to meta-analyse net benefit over a range of risk thresholds.

There are published meta-analyses that compare the performance of different methods to calculate the risk of malignancy in adnexal masses or to classify adnexal masses as benign or malignant but that do not perform head-to head comparisons (46,47). In their meta-analysis that included 47 studies and 19674 adnexal masses, Meys and colleagues (47) assessed the classification performance of subjective assessment (pattern recognition), the IOTA Simple Rules, the IOTA logistic regression model 2, LR2 (10% risk of malignancy cutoff) and RMI (cutoff 200). RMI (meta-analysis of 14 studies) had the poorest classification performance. Its pooled sensitivity was 0.75 (95% CI 0.72-0.79) and its pooled specificity 0.92 (0.88-0.94), i.e. the sensitivity of RMI was higher than in our study (0.75 vs 0.61), while the specificity was similar (0.92 vs 0.92-0.95). Kaijser and colleagues (46) compared the ability of 19 methods to classify adnexal masses as benign or malignant, including the IOTA Simple Rules, LR2 (10% risk of malignancy cutoff) and RMI (200 cutoff). Their meta-analysis included 96 studies and 26438 adnexal masses. In this meta-analysis, too, the classification performance of RMI was poorer than that of the Simple Rules and LR2, and only three of the 19 classification methods performed worse than RMI. The pooled sensitivity of RMI (meta-analysis of 23 studies) was 0.72 (0.67-0.76) and the pooled specificity 0.92 (0.89-0.93), i.e. the results were similar to those of Meys et al (47). The most comprehensive meta-analysis of the diagnostic performance of ADNEX (47 studies, 17007 adnexal masses) reports meta-analysed summary estimates of AUC, sensitivity, specificity and net benefit at the 10% risk of malignancy cutoff (21). The results are very similar to those in the current meta-analysis. Due to lack of information in the studies included in our meta-analysis we could not meta-analyse the net benefit of RMI and ADNEX over a whole range of risk thresholds, nor at different RMI cutoffs. However, in their meta-analysis of data from 17 centres, van Calster al found the net benefit of ADNEX to be higher than that of RMI at cutoff 200 at risk thresholds from 5% to 50% (14). An older, smaller international multicentre study (2403 patients; 18 centres), found the clinical utility of ADNEX to be superior to that of RMI at RMI cutoff 25, 100, 200, 250 and 450 at risk thresholds from 5% to 50%. However, that study used a preliminary version of the ADNEX model (48).

In their article published in 1990, Jacobs et al discuss the choice of cutoff of RMI to decide which patients to refer to oncological care and state that if the availability of such care is limited the cutoff can be set at 75 or 200 (6). Many guidelines have adopted the 200 cutoff (7–12). As shown in this meta-analysis, RMI at 200 cutoff has low sensitivity (0.61) but high specificity (≥0.92). At this cutoff, a high proportion of stage 1 ovarian malignancies, borderline tumors and secondary ovarian metastases will be missed, but most advanced cancers will be detected (49). At the 10% risk of malignancy cutoff of ADNEX, recommended in an international consensus statement (1), also borderline tumors and stage 1 primary invasive ovarian malignancies will be detected and therefore referred to an oncology centre but so will many benign tumors (sensitivity in this meta-analysis ≥0.92, specificity 0.78 - 0.85). This would seem to indicate that lack of gynaecological oncological expertise is no longer a problem (in western countries). This is supported by the recommendation of Sundar et al (12). Sundar et al did a head to head comparison of RMI (cutoff 250) with ADNEX in postmenopausal women with symptoms suggestive of ovarian cancer in a real-world setting (ROCKeTS study), and suggested based on their results that “in view of its higher sensitivity than RMI1 at 250, despite some loss in specificity, we recommend that IOTA ADNEX at 10% should be considered as the new standard-of-care diagnostic in ovarian cancer for postmenopausal patients” (12). This indicates that today, high sensitivity has priority over high specificity. The sensitivity of RMI can be increased by using a lower cutoff, e.g. 25 or 50. However, according to an unpublished meta-analysis of data from 23 Italian centres (submitted for publication; personal communication Antonia Testa), the clinical utility of ADNEX is superior to that of RMI also at RMI cutoffs of 25 and 100 over a whole range of risk thresholds from 1% to 50%.

In conclusion, the results of our meta-analysis support that it is time to consider replacing RMI with ADNEX in clinical guidelines, as suggested by Sundar and colleagues (12). The need for IOTA certification and adherence to a standardised examination technique and terminology may present a challenge but should be seen as a necessary step in the evolution of modern medical practice. Moreover, ADNEX is integrated into ultrasound machines and calculates the likelihood of four different types of malignant tumours. This makes it a valuable tool for clinicians. ADNEX works well also in the hands of ultrasound examiners with limited ultrasound experience (results from an Italian multicentre study submitted for publication, personal communication Antonia Testa) and when used in “a real-world” setting (12).

## ADDITIONAL INFORMATION

### Acknowledgements

We thank Thomas Vandendriessche, Chayenne Van Meel, and Krizia Tuand, the biomedical reference librarians of the KU Leuven Libraries (2Bergen, Learning Centre Désiré Collen, Leuven, Belgium) for their help in conducting the systematic literature search. We gratefully acknowledge the authors of the external validations for providing their valuable data.

### Authors’ contributions

Contributions were based on the CRediT taxonomy. Conceptualization: LB, BVC. Funding acquisition: BVC, DT. Project administration: LB, AL. Supervision: BVC, LV, DT. Methodology: LB, GSC, LW, JYV, BVC. Resources: LB, AL. Investigation: LB, AL, LV, BVC. Validation: LB, AL, PD, GSC, BVC. Data curation: LB, AL, JYV, LV, BVC. Software: LB, AL, LW, BVC. Formal analysis: LB, LW, BVC. Visualization: LB, AL. Writing – original draft: LB, AL, AK, DT, BVC. Writing – review & editing: all authors. All authors have read, share final responsibility for the decision to submit for publication, and agree to be accountable for all aspects of the work.

### Ethics approval and consent to participate

Ethics approval was not required for the systematic review and meta-analysis.

### Consent for publication

Not applicable, we only use already published data.

### Data availability

Data and code to reproduce results and figures are available in a public, open access repository (link https://osf.io/nt89z/). All data relevant to the study are included in the article or uploaded as supplementary information. Some data were provided by the authors and were not public information; therefore, this information was omitted from the public repository.

