## Supplementary Material for "Head-to-head comparison of the RMI and ADNEX models to estimate the risk of ovarian malignancy: systematic review and meta-analysis of external validation studies"

**SUPLEMENTARY MATERIAL**

**Corresponding author**

Ben Van Calster

KU Leuven, Department of Development and Regeneration

Herestraat 49 box 805

3000 Leuven

Belgium

### S1. Search strategy

**Phase 1:**

The following databases were searched for eligible studies:

Search string for **PubMed**: ADNEX [tiab] OR (assessment[tiab] AND "different neoplasias"[tiab] AND "adnexa"[tiab]) AND ("2014/10/18"[Date - Publication] : "3000"[Date - Publication])

Search string for **EMBASE:** ADNEX:ti,ab,kw OR (assessment:ti,ab,kw AND 'different neoplasias':ti,ab,kw AND 'adnexa':ti,ab,kw) AND [2014-2024]/py

Search string for **Web of Science Core Collection**: TS= ("ADNEX" OR ("assessment" AND "different neoplasias" AND "adnexa")) AND PY=(2014-2024)

Search string for **SCOPUS**: TITLE-ABS-KEY (“ADNEX" OR ( "assessment" AND "different neoplasias" AND "adnexa" ) ) AND PUBYEAR > 2013

Search string for **EuropePMC**: adnex AND (SRC:PPR)

Additionally all the citations of ADNEX original paper (https://pubmed.ncbi.nlm.nih.gov/25320247/ ) were retrieved in **PubMed, SCOPUS/EMBASE and Web of Science**. We also searched for studies in systematic reviews that mention the ADNEX model.

**Phase 2:**

For the included articles in phase 1, all the relevant citations and references not already checked in phase 1 were checked in **PubMed, SCOPUS/EMBASE and Web of Science** for inclusion assessment. To determine if a referenced paper or citation was relevant to the systematic review, the title and context in the included paper were used as recommended by Wohlin(1). Phase 2 was performed at data extraction for included papers in phase 1.

The search was first conducted in 29$^{th}$ November 2022 and repeated in 3$^{rd}$ March 2023 , 15$^{th}$ May 2023 and 31$^{st}$ July 2024.

One article was written in a language that was not understood by any of the co-authors (Turkish). Because we did not find suitable translators, we used the automatic translation tool deepl.com.

#### S2. Meta-analysis methods for AUC, sensitivity, specificity

We first extracted all the performance metrics as they were reported in the studies. Some studies presented confidence intervals or standard errors for the metrics, others did not. We used the following methods to approximate the uncertainty.

Approximation of the standard error for the logit of the AUC was based on Newcombe's method 4 (2) implemented in the ccalc function from the "metamisc" R package (3):

$$SE(logit\ c) \approx \frac{SE(c)}{c(1-c)} \approx \sqrt{\frac{1 + n^* \frac{1-c}{2-c} + \frac{(m^* c)}{1+c}}{mnc(1-c)}},$$

where c is the *c* statistic, *n* the number malignant tumours and *m* the number of benign tumours, and $n^* = m^* = \frac{1}{2}(m+n)$.

Approximation of standard error for sensitivity and specificity was based on Wilson's method (4,5) implemented in madad function from the mada R package (6):

$$CI = \frac{\hat{p} + \frac{z^2}{2n} \pm z\sqrt{\frac{\hat{p}(1-\hat{p})}{n} + \frac{z^2}{4n^2}}}{1 + \frac{z^2}{n}},$$

with $\hat{p}$ as reported sensitivity/specificity, *n* the number of tumours, and $z$ denotes the quantile of the standard normal distribution.

To obtain the summary estimates, we performed a random effects meta-analysis to account for differences between studies. Random effects weights were calculated with inverse variance method $w_k = \frac{1}{s_k^2 - \tau^2}$ with $s_k^2$ as the within study variance and $\tau^2$ as the between study variance. We calculated $\tau^2$ using Restricted Maximum Likelihood ("REML") (7).

Subgroup analysis was performed by selecting from the k studies the studies that were part of the subgroup and conducting the meta-analysis independently from the whole sample, hence we did not use a common $\tau^2$.

Confidence intervals of the random effects meta-analysis are constructed using Sidik-Jonkman Hartung-Knapp method (8,9):

$$Var_{HKSJ} = \frac{\sum w_k (\theta_i - \hat{\theta})^2}{K - 1 \sum w_k}.$$

Prediction intervals for the AUC were calculated using Bayesian methods. We used weak priors based on half Student-t distribution with location m = 0, scale σ = 0.5 and ν degrees of freedom = 3 (3).

$$\tau_{discr} \sim Student(0, 0.5^2, 3)\ T\ [0, 10]$$

Prediction intervals for specificity and sensitivity were calculated under the assumption that the random effects of each study are normally distributed with between study standard deviation ($\tau$) as follows:

$$PI = \hat{\mu} \pm\ t_{k-2}\sqrt{\hat{\tau} + SE(\hat{\mu})^2},$$

with $\hat{\mu}$ as the estimated pooled effect, $t_{k-2}$ is the $100\left(1-\frac{\alpha}{2}\right)$ percentile of the t-student distribution with k-2 degrees of freedom, and k the number of studies in the meta-analysis.

#### S3. Trivariate random effects meta-analysis for Net Benefit

We conducted a random effects meta-analysis of Net Benefit (NB) of ADNEX at the 10% risk of malignancy threshold and of RMI at the 200 cutoff according to the methodology described in (10). Relative Utility (RU) expresses NB as a percentage of the maximum possible utility: RU=1 indicates maximum possible utility, RU=0 means no utility, RU<0 means harm. No utility means that NB of ADNEX/RMI is not higher than NB of treating all patients ($NB_{TA}$) and NB of treating no one ($NB_{TN}$, which is 0 by definition). In that case, ADNEX/RMI is not better than simply assuming that everyone needs treatment or that no one needs treatment without the use of any model. Harm means that NB of ADNEX/RMI is lower than $NB_{TA}$ or $NB_{TN}$. Harm means that you can make better decisions without the model.

NB, $NB_{TA}$, and RU are defined as follows:

$NB = Se * P - w * (1 - Sp) * (1 - P)$,

$NB_{TA} = P - w * (1 - P)$,

$RU = \frac{NB - \max(0, NB_{TA})}{P - \max(0, NB_{TA})}$,

with $Se$ sensitivity, $P$ prevalence of malignancy, $Sp$ specificity, and $w$ the odds of the risk threshold. In our case, $w = 1/9$ because our risk threshold is 10%.

We used weak prior distributions for the sensitivity, specificity and prevalence based on the results reported in the ADNEX/RMI model development study. We used normal priors for sensitivity, specificity and prevalence of malignancy. This assures prior probability distributions bounded by 0 and 1. All other priors were the default priors suggested by Wynants et al (10).

For Markov chain Monte Carlo (MCMC) sampling, we used 1000 samples per chain and a burn-in of 1000, and two chains. This was sufficient for convergence and to have Monte Carlo (MC) error <5% of the standard deviation of the posterior distribution for all parameters of interest.

Note that we implemented the methodology described in (11) after considering a recent erratum

### S4. Supplementary tables

**Table S1 Data extracted from each validation study.**

| | Item | Values |
|---|---|---|
| **General information** | Name of the reviewer | Name |
| | Number of validations (e.g. total study population plus postmenopausal gives two validations) | Number of validations |
| | Unit of study | Patient or tumour |
| | Version of ADNEX | With CA125, without CA125, unclear |
| **Target population and setting** | Single country | Yes or No |
| | Number of countries | Number of countries |
| | Start date and end date of recruitment | Start and end date |
| | Target population | Operated only, or both operated and managed with follow-up |
| **Study description** | Study design | Prospective, retrospective, ambispective, unclear cohort. |
| | Setting | Oncology, Non-oncology, unclear |
| | Recruitment method | Consecutive, probably consecutive, other, unclear |
| | Number of centres | Monocentric or multicentric and number of centres |
| | Inclusion criteria & Exclusion criteria | Listed as in the study |
| | Missing data as exclusion criteria | Yes, No, unclear |
| | Exclusion variables | ADNEX predictors or outcome with missing data that resulted in exclusion of the patient or tumour |
| | Number of excluded patients because of missing data | Number of excluded patients |
| | How are borderline ovarian tumours treated | Benign, malignant, other |
| **Predictors** | Measurement used in the study | Mean, Median, Unclear, Not reported |
| | Variability measure | Standard deviation, Interquartile range, Range, Unclear, Not reported |
| | Age of the population | Age (Variability) |
| | Reported descriptive statistics for the ADNEX predictors | Age; CA125; Family history; Maximal diameter; Solid tissue; Papillary projections; >10 cyst locules; Shadows; Ascites |
| | Reported descriptive statistics of predictors by outcome | Total; Benign; Borderline; Stage I; Stage II-IV; Metastatic; Malignant |
| **Subgroup analysis** | Menopausal data | Yes or No |
| | Conservative follow-up | Yes or No; if yes, duration of follow up |
| **Outcome** | Type | Multinomial or binary |
| | Reference standard | Histology, Other |
| | Sample size and number of malignancies and tumour subtypes | Number |
| | Malignancy rate | Percentage |
| **Analysis (ADNEX)** | Missing data | Number of patients with missing information for any variable |
| | Handling of missing data | Complete case analysis, single imputation, multiple imputation, not reported |
| | Software for missing data | Python, R, Stata, SAS, Not reported |
| | AUC as the sum of two triangles | Yes or no |
| | Performance AUC Benign vs Malignant | AUC (CI 95%) |
| | AUC ROC plot | Yes or No |
| | Pairwise AUC methodology | Conditional risk, other, not reported |
| | Performance pairwise AUC | AUC (CI 95%) for the 10 possible pairs |
| | Performance PDI | PDI |
| | Calibration plot | Yes or no |
| | Calibration intercept and slope | Calibration slope and intercept |
| | Multinomial calibration plot | Yes or No |
| | Risk of malignancy cutoff | Cutoff |
| | Sensitivity/Specificity at all cutoffs[a] | Sensitivity, specificity (CI 95%) |
| | PPV and NPV at 10% | PPV, NPV (CI 95%) |
| | DOR at 10% | DOR |
| | Net benefit | Net benefit |
| | Extra reported metrics | Names of metrics |
| **Analysis (RMI)** | Performance AUC Benign vs Malignant | AUC (CI 95%) |
| | Sensitivity/Specificity at 200 cutoff | Sensitivity, specificity (CI 95%) |
| **General information** | Statistical software used | Name of software(s) |
| **TRIPOD** | All applicable tripod items (See Table S2) | Yes or No, if NO with an explanation for our classifying an item as not having been addressed by the authors |
| **PROBAST** | All applicable signalling questions and risk of bias (ROB) assessment by subdomain and overall ROB | Yes, Probably Yes, No, Probably no, No information for signalling questions.<br>Low, Unclear, High ROB<br>Arguments for ROB classification when needed |

AUC, area under the receiver operating characteristic curve; ROC, receiver operating characteristic curve; PDI, polytomous discrimination index; PPV, positive predictive value; NPV, negative predictive value; DOR, diagnostic odds ratio; CI, confidence interval.

[a] These metrics were extracted both for RMI and ADNEX.

**Table S2 Transparent reporting of a multivariable prediction model for individual prognosis or diagnosis (TRIPOD) items.**

| Section/Topic | Item | Checklist Item |
|---|---|---|
| Title | 1 | Identify the study as developing and/or validating a multivariable prediction model, the target population, and the outcome to be predicted. |
| Abstract | 2 | Provide a summary of objectives, study design, setting, participants, sample size, predictors, outcome, statistical analysis, results, and conclusions. |
| Background and objectives | 3a | Explain the medical context (including whether diagnostic or prognostic) and rationale for developing or validating the multivariable prediction model, including references to existing models. |
| | 3b | Specify the objectives, including whether the study describes the development or validation of the model or both. |
| Source of data | 4a | Describe the study design or source of data (e.g., randomized trial, cohort, or registry data), separately for the development and validation data sets, if applicable. |
| | 4b | Specify the key study dates, including start of accrual; end of accrual; and, if applicable, end of follow-up. |
| Participants | 5a | Specify key elements of the study setting (e.g., primary care, secondary care, general population) including number and location of centres. |
| | 5b | Describe eligibility criteria for participants. |
| | 5c | Give details of treatments received, if relevant. |
| Outcome | 6a | Clearly define the outcome that is predicted by the prediction model, including how and when assessed. |
| | 6b | Report any actions to blind assessment of the outcome to be predicted. |
| Predictors | 7a | Clearly define all predictors used in developing or validating the multivariable prediction model, including how and when they were measured. |
| | 7b | Report any actions to blind assessment of predictors for the outcome and other predictors. |
| Sample size | 8 | Explain how the study size was arrived at. |
| Missing data | 9 | Describe how missing data were handled (e.g., complete-case analysis, single imputation, multiple imputation) with details of any imputation method. |
| Statistical analysis methods | 10c | For validation, describe how the predictions were calculated. |
| | 10d | Specify all measures used to assess model performance and, if relevant, to compare multiple models. |
| Risk groups | 11 | Provide details on how risk groups were created, if done. |
| Development vs. validation | 12 | For validation, identify any differences from the development data in setting, eligibility criteria, outcome, and predictors. |
| Participants | 13a | Describe the flow of participants through the study, including the number of participants with and without the outcome and, if applicable, a summary of the follow-up time. A diagram may be helpful. |
| | 13b | Describe the characteristics of the participants (basic demographics, clinical features, available predictors), including the number of participants with missing data for predictors and outcome. |
| | 13c | For validation, show a comparison with the development data of the distribution of important variables (demographics, predictors and outcome). |
| Model performance | 16 | Report performance measures (with CIs) for the prediction model. |
| Limitations | 18 | Discuss any limitations of the study (such as nonrepresentative sample, few events per predictor, missing data). |
| Interpretation | 19a | For validation, discuss the results with reference to performance in the development data, and any other validation data. |
| | 19b | Give an overall interpretation of the results, considering objectives, limitations, results from similar studies, and other relevant evidence. |
| Implications | 20 | Discuss the potential clinical use of the model and implications for future research. |
| Supplementary information | 21 | Provide information about the availability of supplementary resources, such as study protocol, Web calculator, and data sets. |
| Funding | 22 | Give the source of funding and the role of the funders for the present study. |

For more information see (12,13)

**Table S3 Descriptive characteristics of included studies (n=11).**

| Study | Region | Type of centre | Study design | Number of centres | Unit | Clin/Histo Focus[1] | N (benign -malignant) | ADNEX version |
|---|---|---|---|---|---|---|---|---|
| **Diaz (2017) (14)** | South America | Oncology | Retrospective | 1 | Patient | No | 227 (159-68) | Both |
| **Meys (2017) (15)** [2] | Europe | Oncology | Unclear | 1 | Patient | No | 326 (211-115) | With CA125 |
| **Sandal (2018) (16)** | Asia | Oncology | Retrospective | 1 | Patient | No | 191 (138-53) | With CA125 |
| **Szubert (2020) (17)** | Europe | Oncology | Unclear | 2 | Tumor | Yes | 451 (250-201) | With CA125 |
| **Tug (2020) (18)** | Asia | Unclear | Retrospective | 1 | Patient | No | 285 (259-26) | With CA125 |
| **Van Calster (2020) (19)** [2] | Europe | Both | Prospective | 17 | Patient | No | 4905 (3864-1041) | Both |
| **Qian (2021) (20)** | Asia | Oncology | Prospective | 1 | Patient | No | 486 (366-120) | Both |
| **Behnamfar (2022) (21)** | Asia | Oncology | Unclear | 2 | Tumor | No | 284 (260-24) | With CA125 |
| **Oun (2023) (22)** | Asia | Oncology | Prospective | 1 | Patient | No | 100 (71-29) | With CA125 |
| **Wang And Yang (2023) (23)** | Asia | Unclear | Retrospective | 1 | Patient | No | 445 (265-180) | With CA125 |
| **Borges (2024) (24)** | Europe | Oncology | Prospective | 3 | Patient | No | 571 (428-143) | Both |

[1]A study was considered to have clinical or histological focus if the study sample consisted of selected histologies (e.g. only borderline tumours), or a selected subgroup of patients (e.g. only pregnant patients).
[2]Papers extracted by authors PD and GSC

**Table S41. Reported performance for distinguishing benign from malignant tumours (15 validations of ADNEX and 11 validations of RMI). Sensitivity and specificity at 10% cutoff for ADNEX and 200 for RMI.**

| Study | Model | Missing Data handling | AUC Benign versus Malignant | Sensitivity (95% CI) | Specificity (95% CI) | TRIPOD items | Risk of bias |
|---|---|---|---|---|---|---|---|
| Diaz (2017)[a,b] | ADNEX with CA125 | CCA | 0.93 (0.90-0.96) | 92.64 | 83.64 | 61% | High |
| Diaz (2017)[a,b] | ADNEX without CA125 | CCA | 0.925 (0.89-0.96) | 91.17 | 79.87 | 64% | High |
| Diaz (2017)[a,b] | RMI | CCA | 0.875 (0.82-0.93) | 51.47 | 96.85 | 61% | High |
| Meys (2017) [b] | ADNEX with CA125 | MI | 0.93 (0.89-0.95) | 98 (93-100) | 62 (55-68) | 64% | High |
| Meys (2017) [b] | RMI | MI | 0.85 (0.81-0.89) | 71 (62-79) | 79 (72-84) | 61% | High |
| Sandal (2018) [b] | ADNEX with CA125 | CCA | NR | 96.2 (87 - 99.5) | 63.7 (55.2- 71.7) | 50% | High |
| Sandal (2018) [b] | RMI | CCA | NR | 66 (51.7-78.5) | 87.7 (81-92.7) | 46% | High |
| Szubert (2020) | ADNEX with CA125 | CCA | NR | NR | NR | 54% | High |
| Szubert (2020) | RMI | CCA | NR | NR | NR | 50% | High |
| Tug (2020)[a,b] | ADNEX with CA125 | CCA | 0.941 (0.042 ) | 88.5 | 84.6 | 64% | High |
| Tug (2020)[a,b] | RMI | CCA | 0.582 (0.064) | 26.9 | 89.6 | 61% | High |
| Van Calster (2020)[a,b] | ADNEX with CA125 | MI | 0.94 (0.92-0.96) | 91.2 (84.8-95.1) | 85.3 (80.9-88.8) | 100% | Low |
| Van Calster (2020)[a,b] | ADNEX without CA125 | MI | 0.94 (0.91-0.95) | 91.1 (84.5-95.1) | 84.5 (80.1-88.0) | 100% | Low |
| Van Calster (2020)[a,b] | RMI | MI | 0.89 (0.85-0.92) | 60.4 (53.7-66.8) | 95.3 (92.8-96.9) | 96% | Low |
| Qian (2021)[a,b] | ADNEX with CA125 | CCA | 0.94 (0.92-0.96) | 93 (87-97) | 76 (72-81) | 68% | High |
| Qian (2021)[a,b] | ADNEX without CA125 | CCA | 0.94 (0.91-0.96) | 93 (87-97) | 74 (69-79) | 68% | High |
| Qian (2021)[a,b] | RMI | CCA | 0.87 (0.83-0.90) | 55 (46-64) | 93 (90-96) | 64% | High |
| Behnamfar (2022)[a] | ADNEX with CA125 | NR | 0.75 (0.69-0.80) | NR | NR | 46% | High |
| Behnamfar (2022)[a] | RMI | NR | 0.52 (0.46-0.58) | 83.3 (62.6-95.3) | 29.6 (24.1-35.6) | 46% | High |
| Oun (2024) | ADNEX with CA125 | CCA | NR | NR | NR | 43% | High |
| Oun (2024) | RMI | CCA | NR | 0.58 | 0.91 | 46% | High |
| Wang And Yang (2023) [c] | ADNEX with CA125 | CCA | 0.92 (0.90-0.95) | 94.4 (90.0-97.3) | 90.6 (86.4-93.8) | 50% | High |
| Wang And Yang (2023) [c] | RMI | CCA | 0.81 (0.78-0.85) | 70.6 (66.3-77.1) | 92.5 (88.6-95.3) | 50% | High |
| Borges (2024) [a,b] | ADNEX with CA125 | SI | 0.96 (0.94-0.98) | 95.8 (91.1-98.4) | 82.5 (78.5-86.0) | 96% | Unclear |
| Borges (2024) [a,b] | ADNEX without CA125 | SI | 0.96 (0.94-0.98) | 98.6 (95.0-99.8) | 79.7 (75.5-83.4) | 89% | High |
| Borges (2024) [a,b] | RMI | SI | 0.913 | 58.7 | 58.7 | 82% | High |

NR, not reported; Unc, unclear; CCA, complete case analysis; SI, single imputation; MI, multiple imputation; Calibr, calibration; ADNEX, Assessment of Different NEoplasias in the adneXa; RMI, Risk of malignancy index; AUC, Area under the receiver operating characteristic curve; RoB, Risk of bias; TRIPOD, Transparent reporting of a multivariable prediction model for individual prognosis or diagnosis

[a] Included in meta-analysis for AUC in operated patients.

[b] Included in meta-analysis of sensitivity and specificity .

[c] Not included in meta-analysis for AUC because they presented an AUC after dichotomising or categorising risks, i.e. the ROC curve has only 1 point.

### S5 Supplementary figures

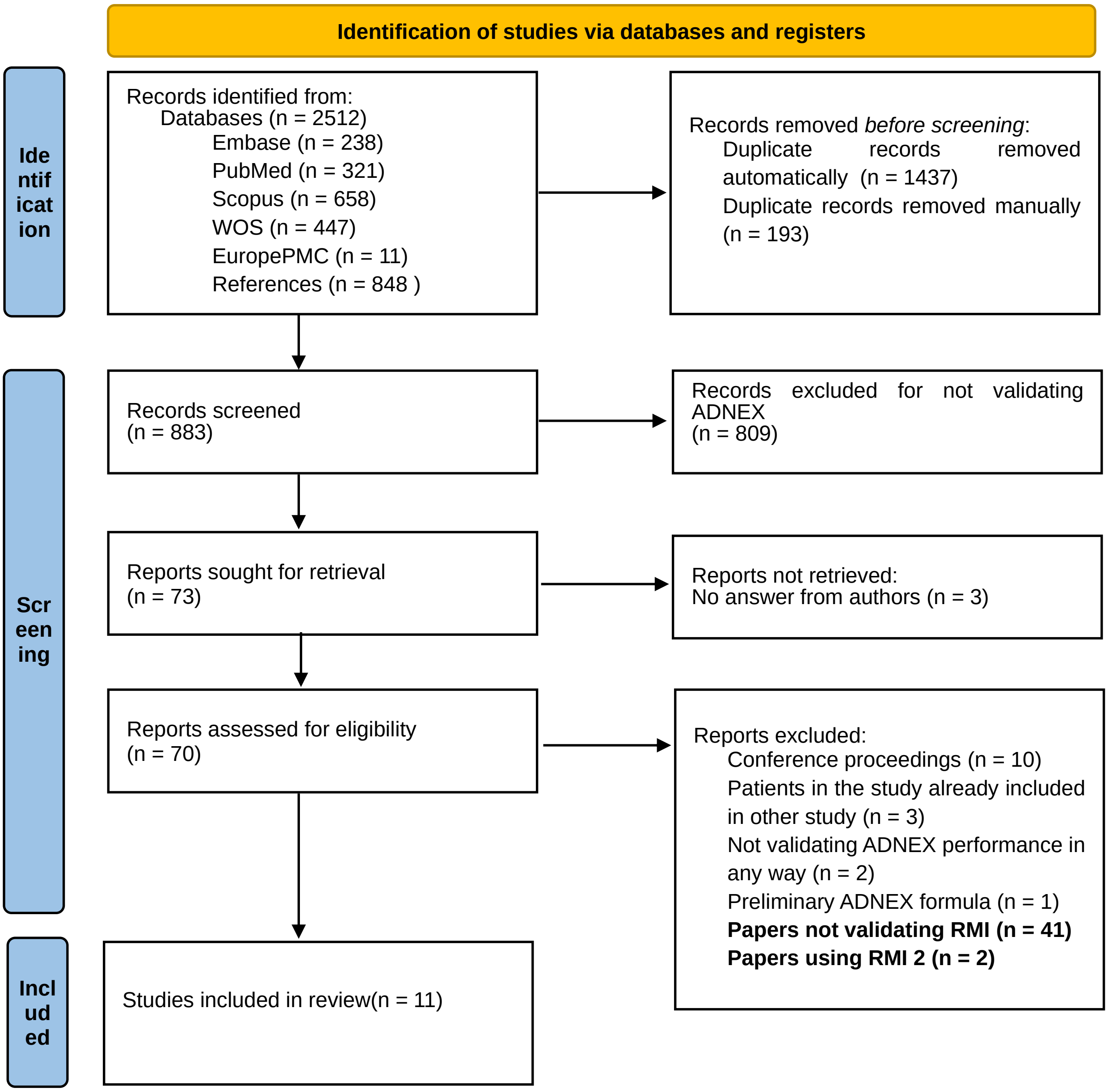


**Fig. S1: PRISMA (preferred reporting items for systematic reviews and meta-analyses) flowchart of study inclusions and exclusions**

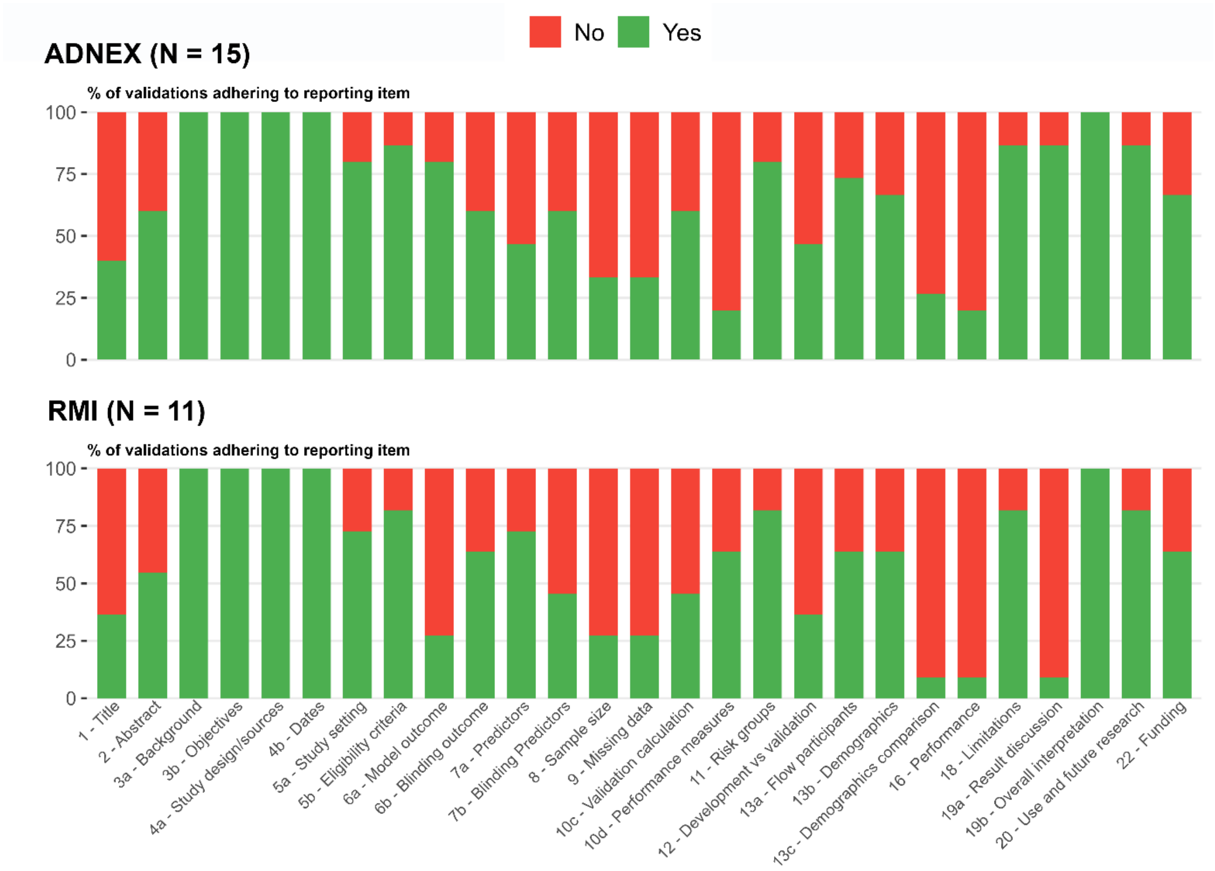


**Fig. S2: Transparent reporting of a multivariable prediction model for individual prognosis or diagnosis (TRIPOD)adherence per item for ADNEX, and RMI. ADNEX, Assessment of Different NEoplasias in the adneXa; RMI, Risk of malignancy index**

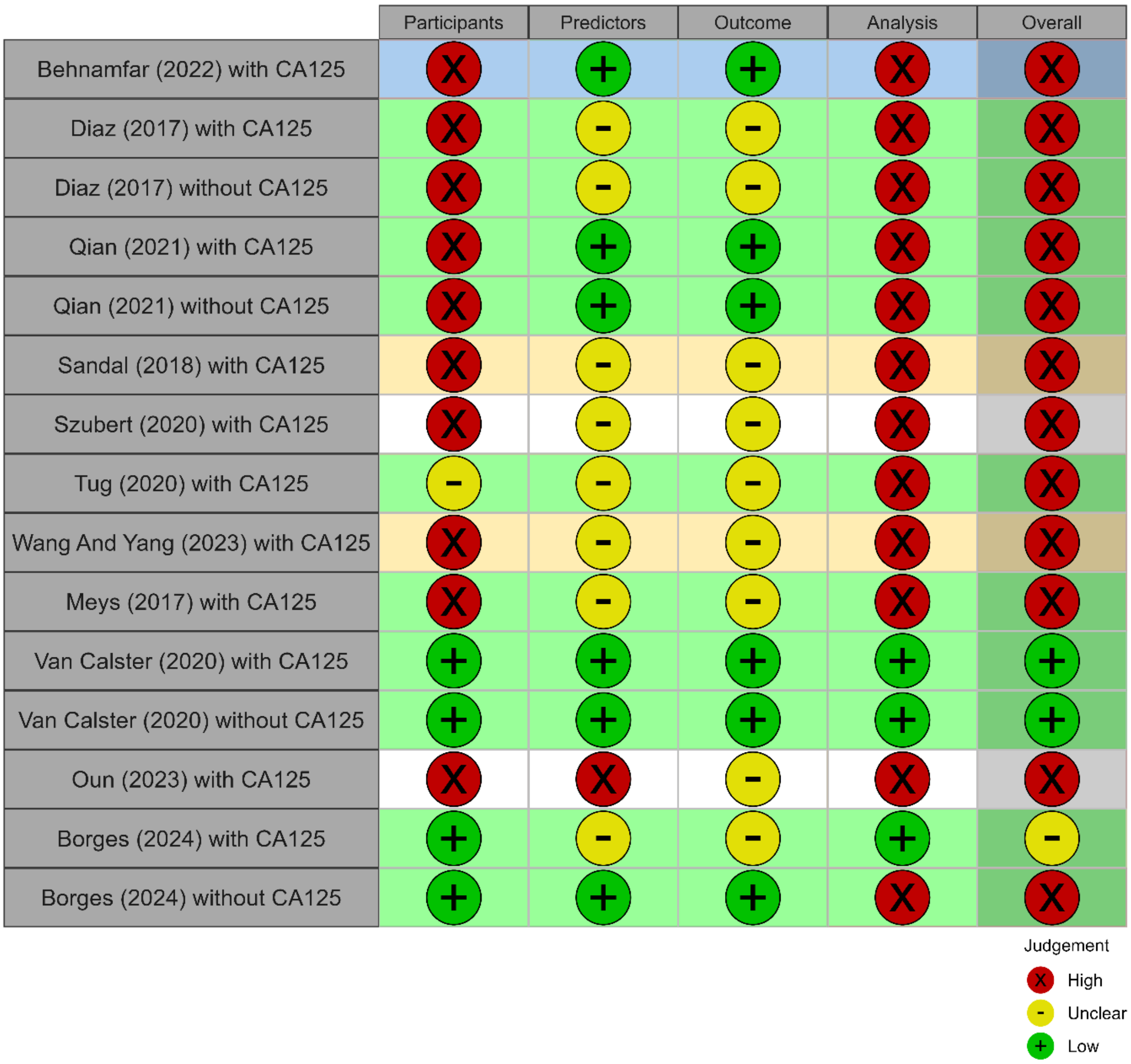


**Fig, S3: PROBAST (Prediction model study Risk Of Bias ASsessment Tool) results by subdomain and overall for studies evaluating Assessment of Different NEoplasias in the adneXa (ADNEX)with and without CA125. Figure generated adapting code from** (25)**. Yellow rows refer to studies that are included in meta-analysis for specificity, sensitivity and clinical utility, blue rows refer to studies included in meta-analysis for the area under the receiver operating characteristic curve (AUC), green rows refer to studies that are included in all meta-analyses, and white rows refer to studies excluded from meta-analysis.**

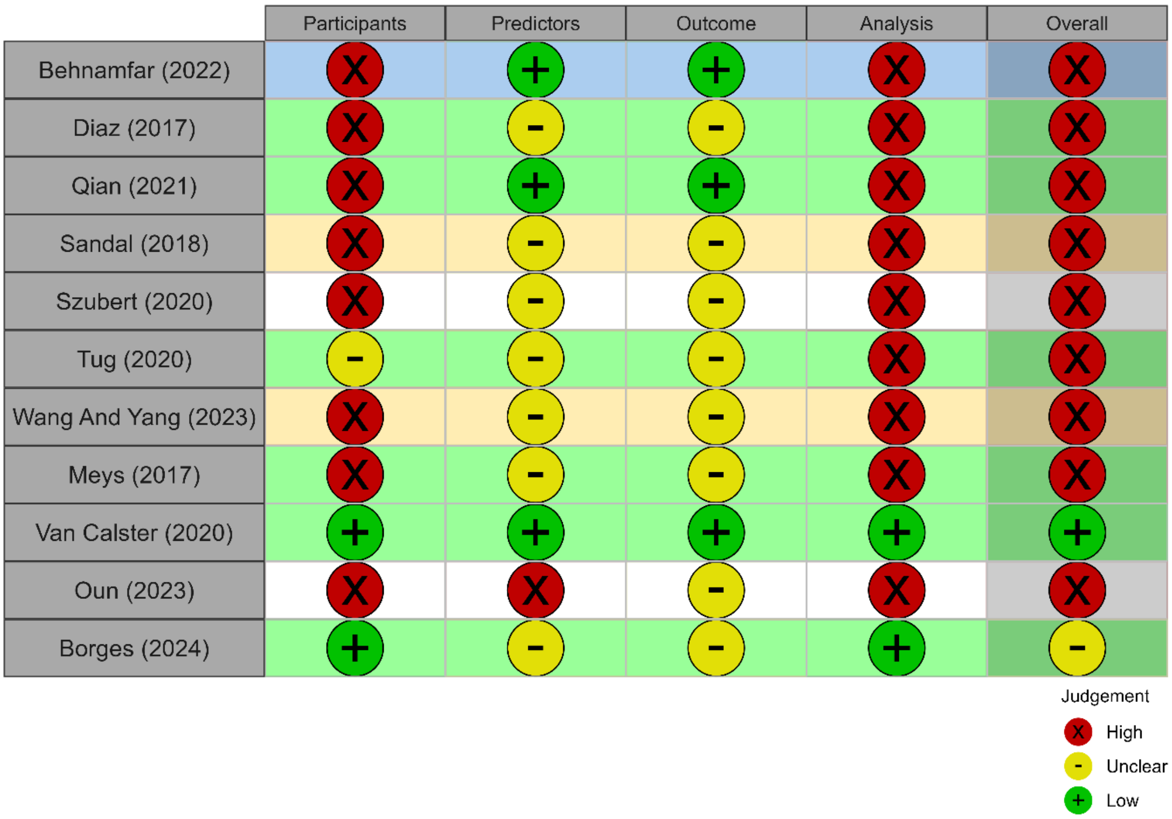


**Fig. S4: PROBAST (Prediction model study Risk Of Bias Assessment Tool) results by subdomain and overall for studies evaluating the Risk of Malignancy Index, RMI. Figure generated adapting code from** (25)**. Yellow rows refer to studies that are included in meta-analysis for specificity, sensitivity and clinical utility, blue rows refer to studies included in meta-analysis for the are under the receiver operating characteristic curve (AUC), green rows refer to studies that are included in all meta-analyses, and white rows refer to studies excluded from meta-analysis.**

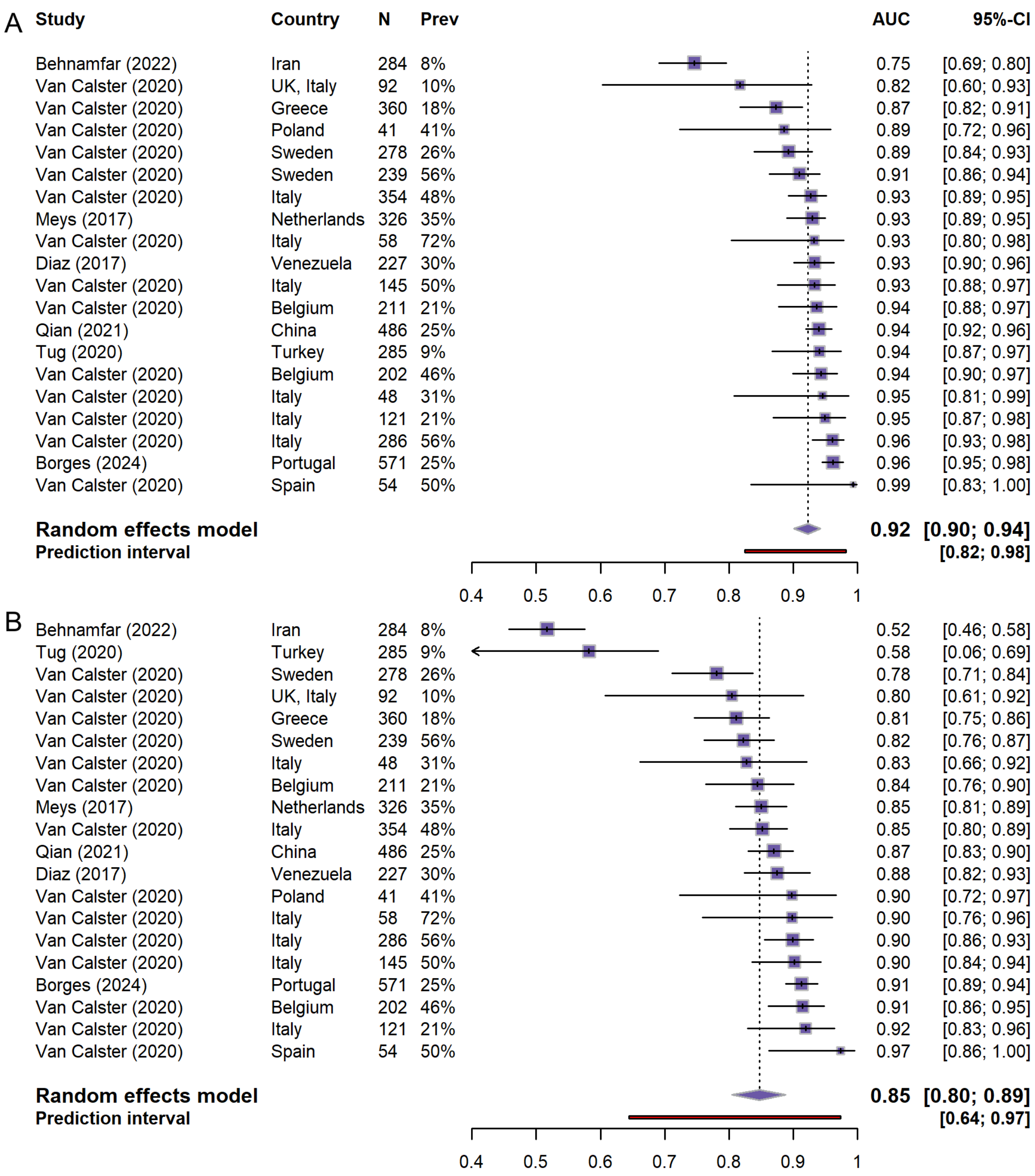


**Fig. S5: Meta-analysis of area under the receiver operating characteristic curve (AUC) of Assessment of Different NEoplasias in the adneXa (ADNEX) with CA125 (A) and Risk of Malignancy Index (RMI) (B) in patients managed surgically. Prev, prevalence; CI, confidence interval.**

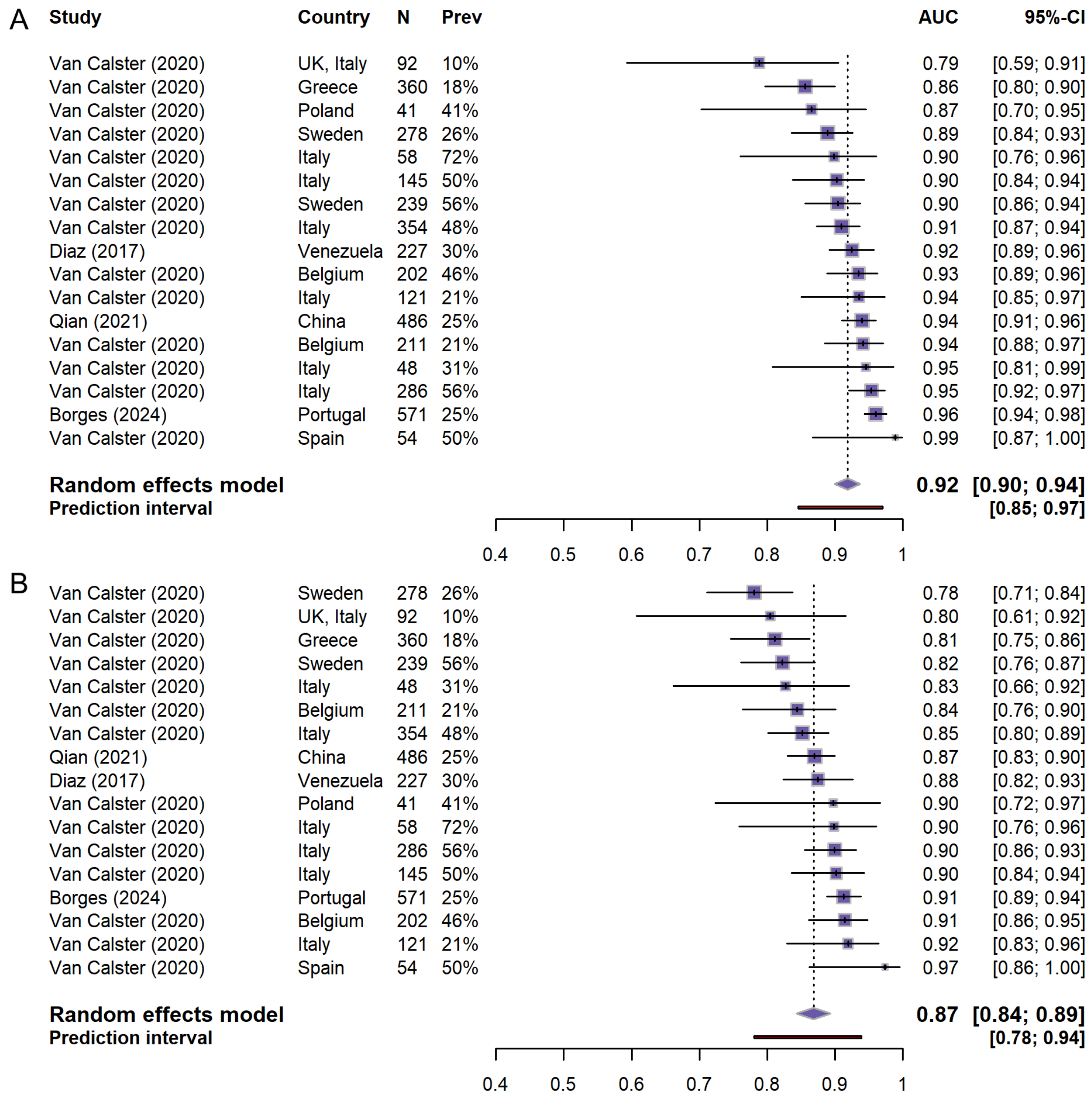


**Fig. S6: Meta-analysis of area under the receiver operating characteristic curve (AUC) of Assessment of Different NEoplasias in the adneXa (ADNEX)without CA125 (A) and Risk of Malignancy Index (RMI) (B) in patients managed surgically. Prev, prevalence; CI, confidence interval.**

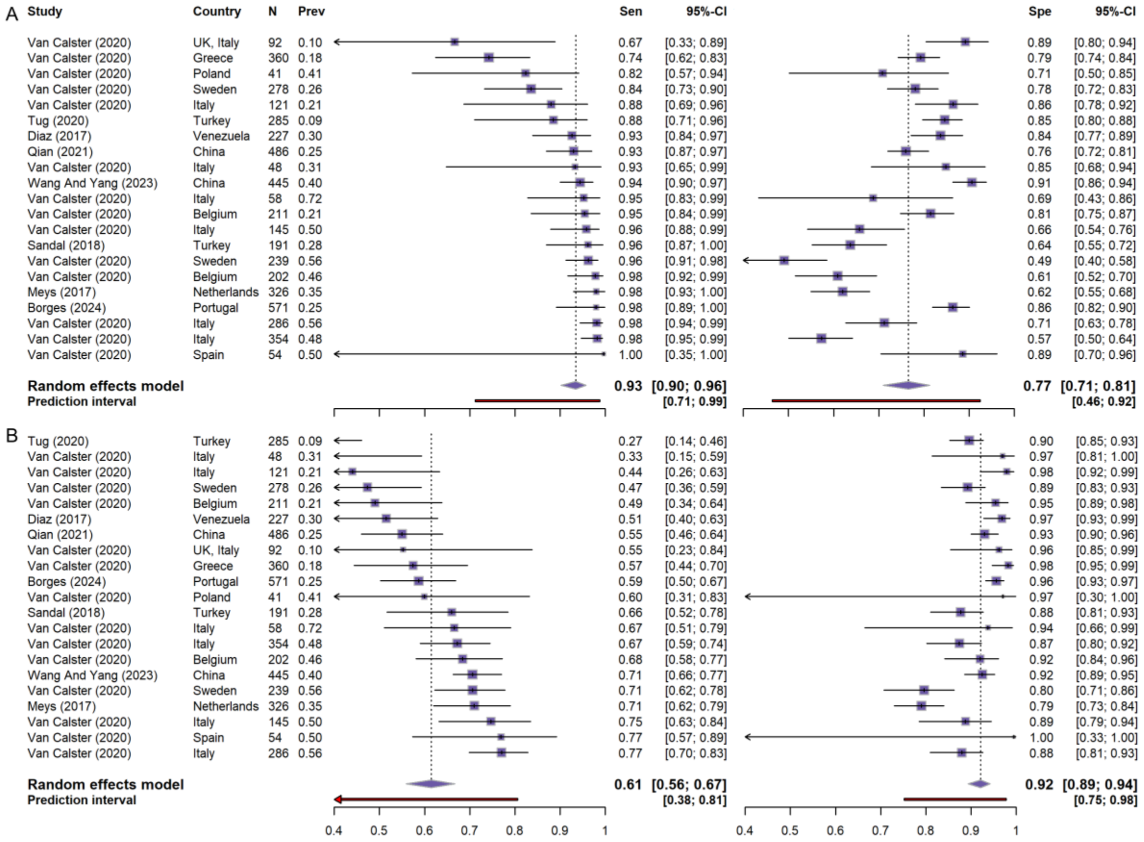


**Fig. S7: Meta-analysis of sensitivity and specificity for Assessment of Different NEoplasias in the adneXa (ADNEX)with CA125 (A) and Risk of Malignancy Index (RMI) (B) in patients managed surgically. Prev, prevalence; CI, confidence interval; Spe, specificity; Sen, sensitivity.**

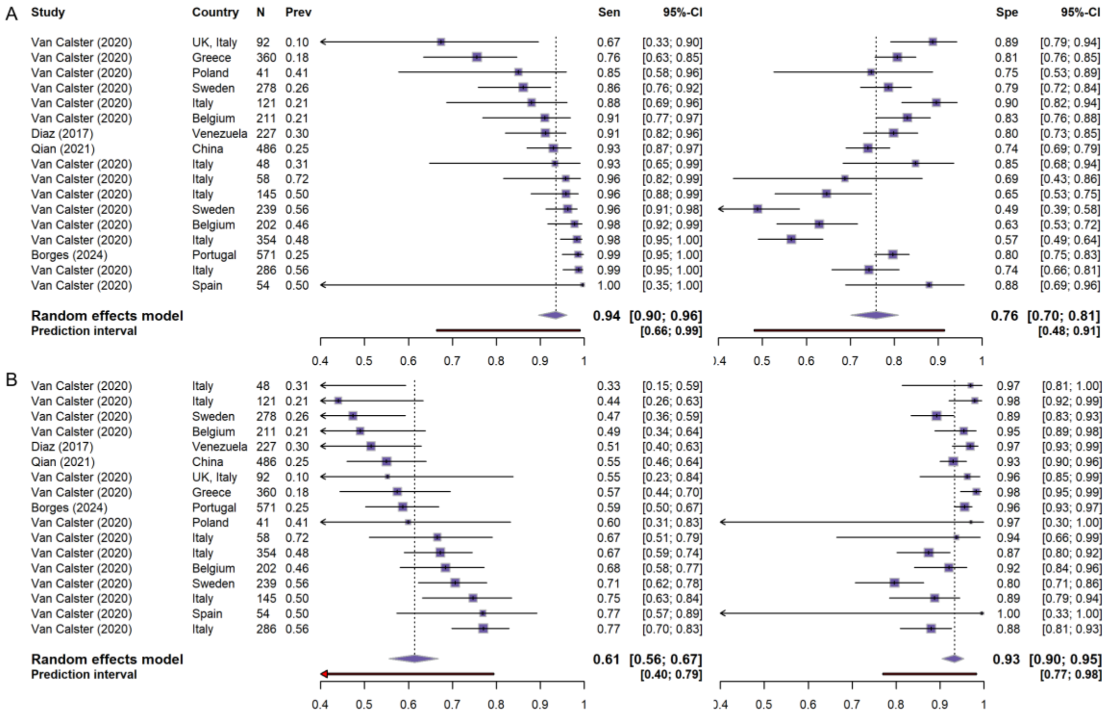


**Fig. S8: Meta-analysis of sensitivity and specificity for Assessment of Different NEoplasias in the adneXa (ADNEX)without CA125 (A) and Risk Of Malignancy Index (RMI) (B) in patients managed surgically. Prev, prevalence; CI, confidence interval; Sen, sensitivity; Spe, specificity.**
